# Efficacy of Nitazoxanide in reducing the viral load in COVID-19 patients. Randomized, placebo-controlled, single-blinded, parallel group, pilot study.

**DOI:** 10.1101/2021.03.03.21252509

**Authors:** Marcelo Silva, Andrés Espejo, María L Pereyra, Martín Lynch, Marcos Thompson, Hernán Taconelli, Patricia Baré, Matías J Pereson, Marcelo Garbini, Pablo Crucci, Diego Enriquez

**Author notes:** **Corresponding author:** Prof. Marcelo Silva, MD. Director de la Unidad de Hepatología y Trasplante Hepático, Hospital Universitario Austral, Pilar, Argentina Av. J. D. Perón 1500 (1629). Derqui, Provincia de Buenos Aires, Argentina.

## Abstract

The fast spread of COVID-19 has overcrowded Public Health Systems facilities in major countries due to the large number of seriously ill patients, particularly those requiring admission to intensive care units. Reducing viral load, along with other recommended epidemiological measures, such as social distancing and home confinement, can in time significantly help to reduce the infection R0 (Basic Reproductive Rate) and then mitigate disease burden. Early negativization or otherwise reduction of the viral load can potentially diminish disease severity, resulting in a better-controlled public health response, avoiding collapse of healthcare systems. Nitazoxanide, a widely used thiazolide approved by the FDA as an antiparasitic drug, also approved in Brazil for Norovirus and Rotavirus treatments, has an excellent safety record for a variety of indications. Nitazoxanide exhibits activity *in vitro* against MERS-CoV and other coronaviruses; and a specific antiviral effect (in micro molar doses) against SARS-CoV-2. The objective of this study was to evaluate the efficacy and safety of Nitazoxanide in reducing the SARS-COV 2 viral load within 7 days of treatment in respiratory samples from COVID-19-infected patients with mild to moderate disease, compared to placebo. An interim analysis showed that the ratio of patients with a viral load reduction ≥ 35% from baseline up to day 7 of treatment was significantly greater for Nitazoxanide compared to placebo (47.8% vs. 15.4%; Δ 34.6%; 95% CI: 64.7; 4.6: *p* = 0.037).

**KEY POINTS:** *State of the Art:*

- Different studies conclude that viral load (VL) would correlate with morbidity, mortality and contagiousness of COVID-19.
- Early negativization or reduction of the viral load can potentially reduce the severity of this disease.
- *In vitro* data demonstrated a specific antiviral effect of Nitazoxanide for SARS-CoV-2.

*Article contribution:*

- Nitazoxanide showed a statistically significant difference versus placebo in the number of patients who had their viral load reduced by at least 35% in mild to moderate COVID-19 disease.
- The observed antiviral effect *in vitro* would seems to be verified in patients with mild to moderate COVID-19 infection, which should be confirmed by studies with a larger cohort of patients.

## Introduction

The fast spread of COVID-19 has overcrowded Public Health Systems facilities in major countries due to a large number of seriously ill patients and particularly those requiring intensive care unit admissions. The clinical spectrum of COVID-19 disease is quite variable, with 80% of the cases being asymptomatic or reporting mild symptoms, and about 5-10% presenting with severe forms, including respiratory failure, which may require intensive care unit assistance and mechanical ventilation *(Wang et al., 2020; Cascella et al., 2020)*. Reducing viral load, along with other epidemiological measures, such as social distancing and home confinement, can in time significantly reduce the infection R0 (Basic Reproductive Rate) and help mitigate disease burden.

Symptomatic and supportive treatment has so far been the main intervention for patients with moderate to severe infection. Recent focus has been oriented to the potential benefit of early treatment in patients with severe coronavirus 2 (SARS-CoV-2) infection and high risk for severe outcomes. However, there is a remarkable lack of treatments alternatives with proven efficacy for patients with mild to moderate early infection. The potential benefits of such treatments include improvement in patient outcomes and prevention of hospital admissions. Longer-term outcome benefits may include prevention of chronic sequelae of infection, and of transmission of SARS COV-2 infection by shortening disease length duration.

Outpatient COVID-19 treatments, along with an effective vaccine, would have significant implications in the struggle against this pandemic. Nevertheless, effective treatments for people with mild to moderate disease have hardly been studied. For instance, remdesivir requires daily infusions for up to 10 days, which is not suitable for outpatient settings *(Beigel et al., 2020)*. Similarly, dexamethasone has not been tested in early and mild disease, whereas its immunosuppressive effects could even worsen clinical outcomes in this setting of profuse viral replication. Several drugs, such as hydroxychloroquine, despite their purported positive effect in non-controlled clinical studies, did not prove to be effective in controlled rigorous clinical trials *(Abella et al., 2021)*. Furthermore, the risk-benefit balance is not the same in mild to moderate disease as in severe disease. Outpatient treatments for mild disease should be safe, easily administered, and scalable, and should have few adverse events. Based on this situation, a group of new therapies has now entered the clinical development stage *(Kim et al., 2020)*.

Given the current critical circumstances, new drugs development would take much longer than acceptable time for a proper management of this pandemic. Therefore, drugs already available for other indications are being tested through different techniques for their potential antiviral effect. Thus, a range of drugs has been tested, including: lopinavir-ritonavir, azithromycin, hydroxychloroquine, colchicine, ivermectin, remdesivir, tocilizumab, methylprednisone. The lack of a drug with a demonstrated strong antiviral efficacy warrants exploring new therapeutic options against SARS-CoV-2/COVID-19.

Nitazoxanide (NTX) is a widely used thiazolide, approved by the FDA and other health authorities worldwide as an antiparasitic, with an excellent safety record for a variety of indications. It was subsequently approved in Brazil for the treatment of Norovirus and Rotavirus and was shown to have broad antiviral activity, including animal and human coronaviruses *(Wang et al., 2020; Rossignol, 2014; Hui et al., 2018)*. NTX has also shown *in vitro* activity against MERS-CoV and other coronaviruses, inhibiting the expression of the viral N protein *(Rossignol, 2016)*. A different proposed antiviral mechanism for NTX involves RNA-dependent protein kinase R (PKR) and α-subunit of eukaryotic translation initiation factor 2 (eIF2α) phosphorylation, with subsequent depletion of ATP-intracellular calcium deposits inducing chronic sub-lethal stress in the endoplasmic reticulum and impairing the glycosylation of the structural N protein in infected cells *(Ashiru et al. 2014)*. On the other hand, NTX up-regulates innate antiviral mechanisms by amplifying the detection of cytoplasmic RNA through the activation of type I interferon (IFN-1) pathways *(Jasenosky et al., 2019)*. Similarly, apart from up-regulation of IFN-1 levels, NTX promotes the expression of intracellular genes that induce the antiviral-state in cells *(Hui et al., 2018)*. Thus, NTX positively regulates the innate immune response by inhibiting ongoing viral replication in infected cells and potentially helping to prevent infection of uninfected cells by avoiding horizontal re-infection. Animal studies also showed a marked decrease in interleukin 6 (IL-6) production, one of the main mediators of the cytokine storm observed in respiratory viral infections. Studies in mice showed that, 6 hours after oral administration, NTX suppresses by 90% the production of lipopolysaccharide-induced IL-6, suggesting that it could also help reducing the cytokines storm observed in respiratory viral infections *(Hong et al., 2012; Chen et al., 2010)*. (For a full review of the possible mechanisms of action of Nitazoxanide in COVID-19, see *Lokhande et al., 2021*).

NTX after oral administration is rapidly hydrolyze to its active metabolite Tizoxanide (TZ), which has a half-life of approximately 2 h, a Cmax of 10.6 µg/ml, and a Tmax of 3 h *(Balderas-Acata et al., 2011)*. When not administered with food, the Cmax and AUC are reduce by half. A study which showed important NTX *in vitro* activity suggested a EC50 for SARS-CoV2 of 2.12 µM (0.62 µg/ml); such steep slope suggests a specific antiviral effect, with an EC90 of around 5-6 µM (1.84 µg/ml) *(Wang et al., 2020)*. This was observe for both, NTX and TZ molecules. The remarkably low EC50 seen for all coronaviruses suggest that their interferon-blocking mechanisms are extremely sensitive to NTX. A physiologically based pharmacokinetic (PBPK) study using a validated computational model concluded that doses of NTX 900 mg TID (with food) would optimally maintain plasma and lung drug levels continually above the CE90 for SARS-COV-2 *(Rajoli et al., 2020)*.

A recently published clinical trial in patients with uncomplicated influenza-like illness, showed a significant reduction in the mean time to symptoms relief, from 108 to 94.9 h (*p*=0.0052) *(Haffizulla et al., 2020)*.

In a more recent, multicenter, randomized, double-blind, placebo-controlled study in COVID-19 patients, after 5 days of NTX 500 mg TID treatment, viral load was significantly lower in the NTX when compared with the placebo group (*p*=0.006). Additionally, on treatment day 5, there were significantly more COVID-19 negative patients in the NTX group than in the placebo group (29.9% *versus* 18.2%) (*p*=0.009) *(Rocco et al., 2020)*.

Considering its previously studied antiviral effect, its safety profile and the possibility of oral use in adults and children, as well as its rapid availability and affordability, NTX is a suitable alternative to be studied. Currently, the potential of NTX in the treatment of COVID-19 is being investigated in more than 25 clinical studies around the world *(Clinicaltrials*.*gov)*.

The objective of this study was to evaluate the efficacy and safety of NTX 500 mg QID plus standard of care for 14 days in patients ≥ 18 years of both sexes infected with COVID-19 with mild to moderate symptoms, versus a placebo plus standard-of-care control group. The original protocol included an interim analysis to find any trends upon enrolment of 1/3 of the proposed number of patients. This article presents the results of this interim analysis.

## Materials and methods

This was a randomized, placebo-controlled, single-blinded, parallel-group, pilot study, carried out at Hospital Universitario Austral, and Sanatorio Nuestra Señora del Pilar, Buenos Aires, Argentina *(Clinicaltrials*.*gov, registry number: NCT04463264)*.

Selected subjects included people of both sexes aged ≥ 18 years, with COVID-19, with a positive quantitative real-time polymerase reaction (rtq-PCR), no more than 4 days after the onset of symptoms. Patients with a chest Rx or lung ultrasound consistent with COVID-19 pneumonia were included if they had mild symptoms, defined as SpO_2_ ≥ 95% when breathing room air and a RF < 24 per minute, or moderate symptoms, defined as SpO_2_ ≥ 92% when breathing FiO_2_ through a low flow O_2_ cannula (< 5 l per minute). Pregnant or lactating women and subjects with any contraindication to the use of NTX (according to its package leaflet) were excluded.

Subjects enrolled at research sites were randomized 2:1 to receive NTX orally with food for 14 days, or placebo plus standard supportive symptomatic treatment for a confirmed diagnosis of COVID-19. The protocol began with 1 g every 8 hours (3 g/day), but soon afterwards, it was changed to 500 mg (1 film-coated tablet) every 6 hours (2 g/day), since the first 2 subjects reported gastrointestinal intolerance with the former dose scheme. A randomization table, unknown to the investigator up to the assignment time was generated using the RANDOM software (Randomness and Integrity Services Ltd. Premier Business Centers, 8 Dawson Street, Dublin 2, D02 N767, Ireland).

The study primary endpoint was viral eradication from the patients’ respiratory tract secretions on day 7 after starting treatment (a viral eradication of at least 35% would be clinically relevant), being the secondary endpoint a viral load reduction from respiratory secretions on days 7, 14 and 35 after starting the treatment compared to baseline. Tolerability was assessed by the occurrence of drug-related adverse events, whether spontaneously reported by the patient, or arising during questioning and/or examination by the investigator.

Samples for SARS-CoV-2 genome detection were collected by healthcare staff trained in upper respiratory tract, by means of nasopharyngeal and oropharyngeal swabs, combined in a single tube pre-filled with 1 mL of DNA/RNA Shield™ (DNA/RNA Shield™ Collection Tube w/ Swab, ZYMO Research). The reagent inactivates pathogens and the nucleic acid content of samples is stabilized during storage/transport. The samples were stored in the Hospital Universitario Austral laboratory for no more than 48 hours, in a refrigerator at 4°C (39.2°F), and then carried in viral transport medium to the laboratory of the National Academy of Medicine. Genomic extraction was carried out with the QIAamp Viral RNA minikit (Qiagen) following manufacturer’s instructions. The virus genome was amplified using the CDC RT-qPCR protocol for Wuhan virus (nCoV-19), 2019-nCov CDC USA of IDT. Detection was carried out on the basis of real-time PCR, including a set of oligonucleotides primers and double-marker hydrolysis probes (Taqman®) (2019 nCov_N1 and N2) that amplify regions of the viral nucleocapsid gene (N). An internal control for RNase P (RP) was also included. A standard curve was generated by plotting the Cq values against the log quantity of known starting sample serial dilutions of viral RNA genome copies/ µl of a purified SARS-CoV-2 RNA (AMPLIRUN® SARS-CoV-2 RNA control). A linear regression was performed in order to interpolate the quantity of viral RNA in each of the test samples from the Cq values. Viral loads correspond to log base 10 transformed values from the N1 target.

These values should be interpreted considering that the lower the VL, the higher the N1 Cq, since that would imply a greater number of reaction replication cycles required to detect the VL.

### Statistical analysis

The N1 Cq and N1 VL log values comparing both treatment arms were analyze for days 1 and 7. An analysis of variance with repeated measures using one factor was used. Furthermore, the absolute and relative variation between days 1 and 7 results were also assessed. Differences in treatment arms were assessed by using a *t* test mean difference. As a secondary efficacy endpoint, the same measures were assessed, but considering their variation between day 1, 7 and 14. Finally, the proportion of patients with a variation ≥ 35% in the N1 Cq and N1 VL log values on day 7 and the proportion of patients with a negative VL result were compared (VL <100 is considered a negative result since, although it is not zero, this value corresponds to residual virus or genome remnants that remain detectable). In this case, a proportion difference *t* test was used.

For the statistical analysis, the SPSS software (IBM version 16.0) was used.

Regarding the sample size, in mild SARS COV-2 infections, the median reported duration of the virus in upper respiratory tract samples was reported to be 18 days (IQR 13-29 days) *(Zheng et al., 2020; De Chang, 2020*. We then expect a 50% of the patients to eradicate the virus within a 3 week period, a 25% before day 13 and that less than 15% during the first week of symptoms onset. From an epidemiological point of view, increasing the viral eradication rate from below 15% to over 30% during the first two weeks of treatment would be clinically relevant. Considering an expected proportion of viral eradication of 13% (CI 5.0-26.7) for the placebo group, and 35% (CI 24.7-45.2) for the treatment group, at the end of week 1, a minimum sample size of 135 COVID-infected patients would be required with a single-tailed upper limit (alpha set to 0.025), 80% power, and a randomization ratio of 2:1. This scheme would require 45 subjects for the placebo arm and 90 patients for the NTX arm.

The study was conducted in compliance with the ethical principles established in the Declaration of Helsinki version 2013, the Guidelines for Good Clinical Practice of the International Council for Harmonization (ICH-GCPs) and the current national regulatory standards, Resolution 1480/11 and applicable legislation in Buenos Aires province. The study was approved by the Institutional Evaluation Committee of the Faculty of Biomedical Sciences of the Universidad Austral. All participating subjects gave their informed consent.

## Results

The Enrollment period was from July 2020 to December 2020. A total of 46 patients were randomized, 33 to the NTX arm and 13 to the placebo arm. (Figure 1).

**Figure 1.**
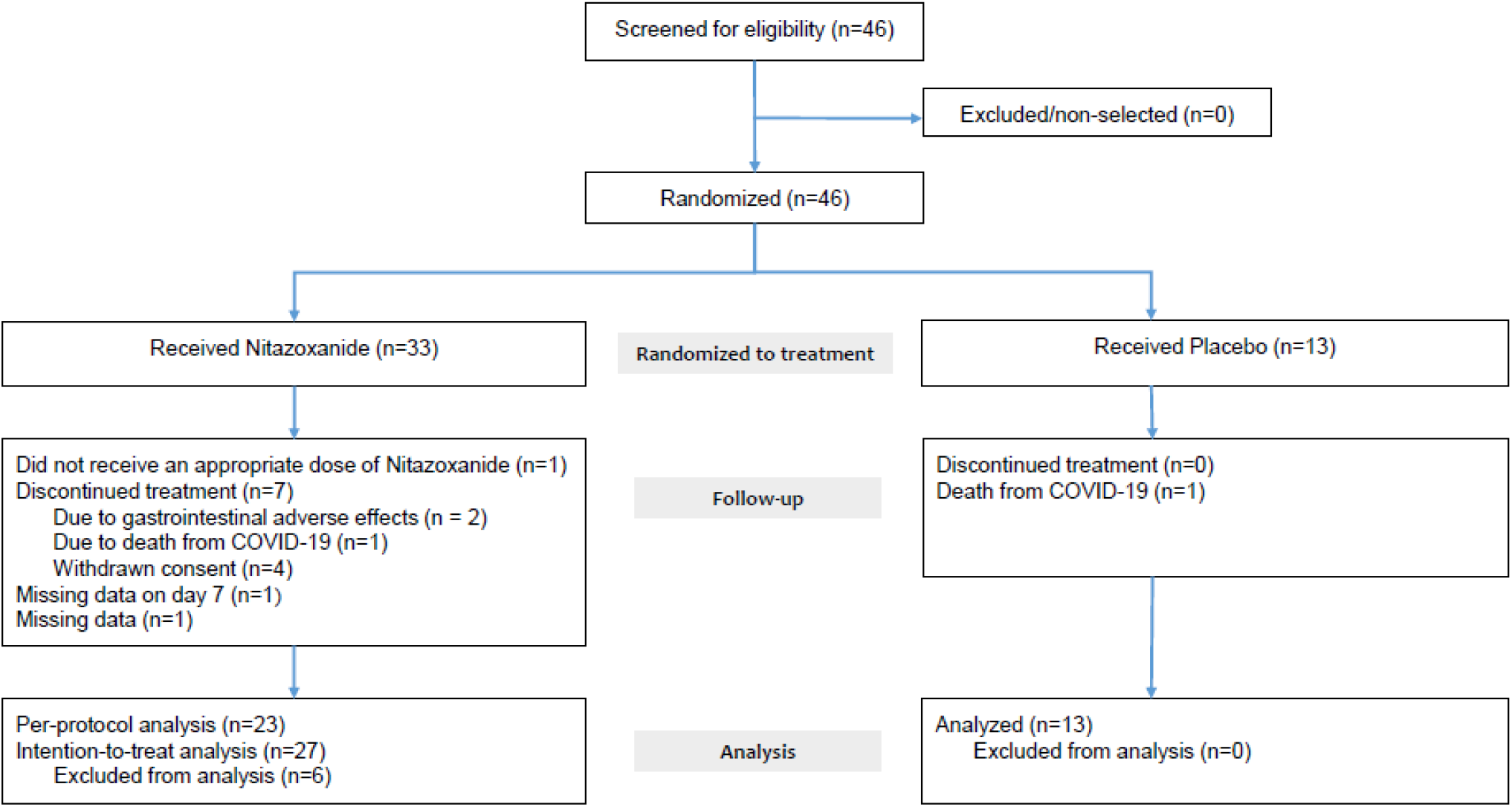
Enrollment.

The demographics and clinical characteristics of the subjects at the beginning of the study were balanced between both groups (Table 1). The median age in the NTX arm was 44 years (19-67) and in the placebo arm it was 53 (28-68). There was a higher proportion of male patients in both groups. Most of the patients were hospitalized with mild disease without additional O_2_ therapy (value 3 on the Ordinal Scale). Three patients treated with NTX reported HR values out of range: 2 patients presented a HR below 60, and the other one had a HR above 100. All patients in the placebo arm showed a HR within the range of 60-100. The patients in both treatment arms had fR values below 20. There was not a significant statistic difference in the medians of lymphocytes, CRP and D-dimer between groups.

**Table 1.**
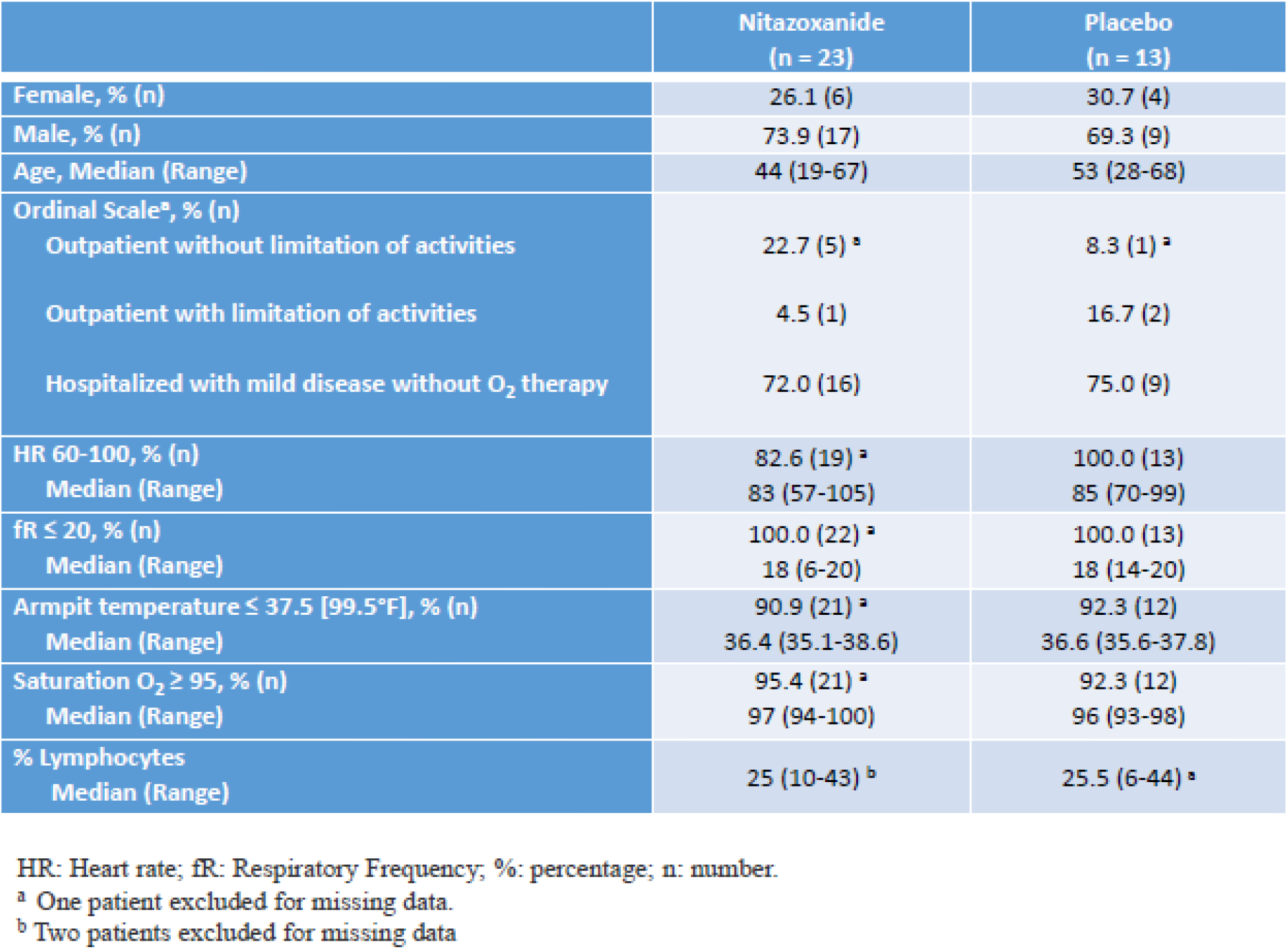
Baseline clinical characteristics

|  | Nitazoxanide<br>(n = 23) | Placebo<br>(n = 13) |
| --- | --- | --- |
| Female, % (n) | 26.1 (6) | 30.7 (4) |
| Male, % (n) | 73.9 (17) | 69.3 (9) |
| Age, Median (Range) | 44 (19-67) | 53 (28-68) |
| Ordinal Scale<sup>a</sup>, % (n) |  |  |
| Outpatient without limitation of activities | 22.7 (5) <sup>a</sup> | 8.3 (1) <sup>a</sup> |
| Outpatient with limitation of activities | 4.5 (1) | 16.7 (2) |
| Hospitalized with mild disease without O<sub>2</sub> therapy | 72.0 (16) | 75.0 (9) |
| HR 60-100, % (n) | 82.6 (19) <sup>a</sup> | 100.0 (13) |
| Median (Range) | 83 (57-105) | 85 (70-99) |
| fR ≤ 20, % (n) | 100.0 (22) <sup>a</sup> | 100.0 (13) |
| Median (Range) | 18 (6-20) | 18 (14-20) |
| Armpit temperature ≤ 37.5 [99.5°F], % (n) | 90.9 (21) <sup>a</sup> | 92.3 (12) |
| Median (Range) | 36.4 (35.1-38.6) | 36.6 (35.6-37.8) |
| Saturation O<sub>2</sub> ≥ 95, % (n) | 95.4 (21) <sup>a</sup> | 92.3 (12) |
| Median (Range) | 97 (94-100) | 96 (93-98) |
| % Lymphocytes |  |  |
| Median (Range) | 25 (10-43) <sup>b</sup> | 25.5 (6-44) <sup>a</sup> |

HR: Heart rate; fR: Respiratory Frequency; %: percentage; n: number.
<sup>a</sup> One patient excluded for missing data.
<sup>b</sup> Two patients excluded for missing data

The primary and secondary efficacy analysis were performed as per protocol. It includes patients with complete data on days 1 and 7 and who received the indicated treatment (two patients without data on day 7 and one patient randomized to NTX but treated with doses well below the protocol doses were excluded). In total, 36 patients were included in this analysis, 23 into the Nitazoxanide arm and 13 into the placebo arm.

Table 2 shows that both treatment arms presented a statistically significant decrease in the viral load (increased N1 Cq) between days 1 and 7, (F = 63.053; *p*< 0.001); this increase is greater in the NTX arm. However, there is no statistically significant difference when comparing N1 Cq increases (F= 1.552; *p*= 0.221) between treatment arms. Thus, the N1 Cq variation is significant, notwithstanding treatment, although the NTX arm tended to show a greater variation.

**Table 2.** Primary efficacy results (N1 Cq) per treatment arm at day 1 and 7. Mean and standard deviation. Per-protocal analysis.

| N1 Cq | Nitazoxanide<br>n=23 |  | Placebo<br>n=13 |  | Total<br>n=36 |  |
| --- | --- | --- | --- | --- | --- | --- |
|  | Mean | SD | Mean | SD | Mean | SD |
| Day 1 | 23.3 | 5.5 | 25.0 | 5.8 | 24.0 | 5.6 |
| Day 7 | 30.8 | 3.9 | 30.5 | 4.4 | 30.7 | 4.0 |

Day 1-7; $F=63.053$ ; $p<0.001$
Day 1-7\*Treatment; $F=1.552$ ; $p=0.221$

Table 3 shows similar values for the absolute and relative variation in N1 Cq between Day 1 and Day 7 results. The difference between treatment arms for absolute and relative variation is not statistically significant (*p* = 0.221 y *p* = 0.358 respectively). However, variations show to be greater for the NTX arm (7.5 vs. 5.5 absolute difference and 37.4 vs. 27.5 relative difference).

**Table 3.** Primary efficacy results (absolute and relative variation in N1 Cq between Day 1 and Day 7) by treatment arm. Mean and standard deviation. Per-protocal analysis

| N1 Cq | Nitazoxanide<br>n=23 |  | Placebo<br>n=13 |  | Dif. | p | 95% CI of the difference |  |
| --- | --- | --- | --- | --- | --- | --- | --- | --- |
|  | Mean | SD | Mean | SD |  |  |  |  |
| Abs. Variation | 7.5 | 4.6 | 5.5 | 5.0 | 2.0 | 0.221 | -1.3 | 5.4 |
| Rel. variation | 37.4 | 28.6 | 27.5 | 34.7 | 9.9 | 0.358 | -11.8 | 31.8 |

Table 4 shows that 47.8% of NTX-treated patients achieved a viral load reduction (increased N1 Cq) ≥ 35%, compared to 15.4% of placebo patients. The difference (32.4%, 95% CI; 2.1, 62.8) is statistically significant (*t* = 2.178; p = 0.037), the ratio of patients with a variation ≥ 35% being significantly higher in the NTX arm.

**Table 4.**
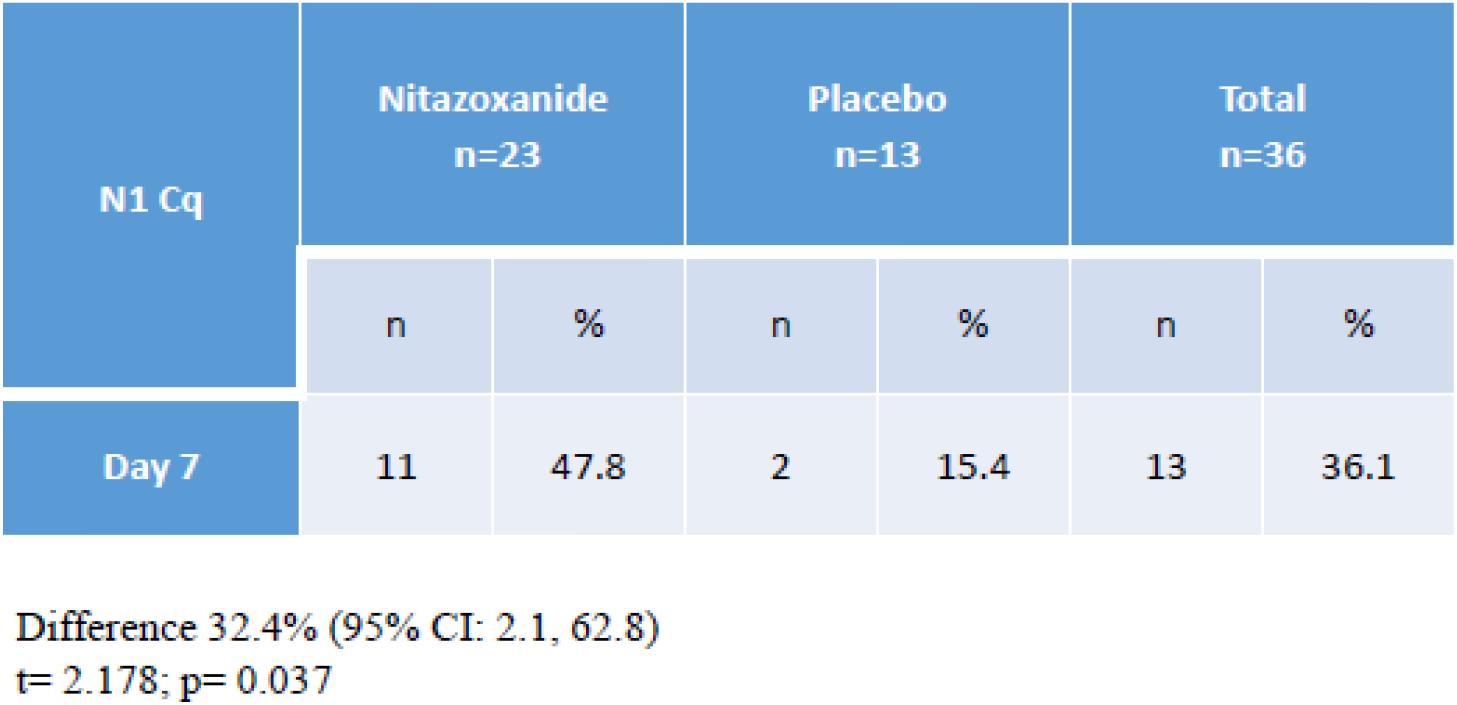
Secondary efficacy results (Patients with a variation ≥35% in N1 Cq) by treatment arm. Per-protocal analysis

| N1 Cq | Nitazoxanide<br>n=23 |  | Placebo<br>n=13 |  | Total<br>n=36 |  |
| --- | --- | --- | --- | --- | --- | --- |
|  | n | % | n | % | n | % |
| Day 7 | 11 | 47.8 | 2 | 15.4 | 13 | 36.1 |

Difference 32.4% (95% CI: 2.1, 62.8) t= 2.178; p= 0.037

The results corresponding to N1 VL log per treatment arm, absolute and relative variation and numsber of patient with a variation ≥35% in VL, on day 1 and 7, are attached in Annex 1.

Graph 1 shows the response-to-treatment assessment, based on the viral load (VL) negativization results on day 7 (VL values <100 copies). 62.5% patients in the NTX arm and 53.9% patients in the placebo arm were negative on day 7. The difference between negativization ratios (8.7%) is not statistically significant (t = 0.500; *p* = 0.620); however, the results tend to favor NTX. The VL log transformation values are similar to the above results (data available for review on file).

**Graph 1.**
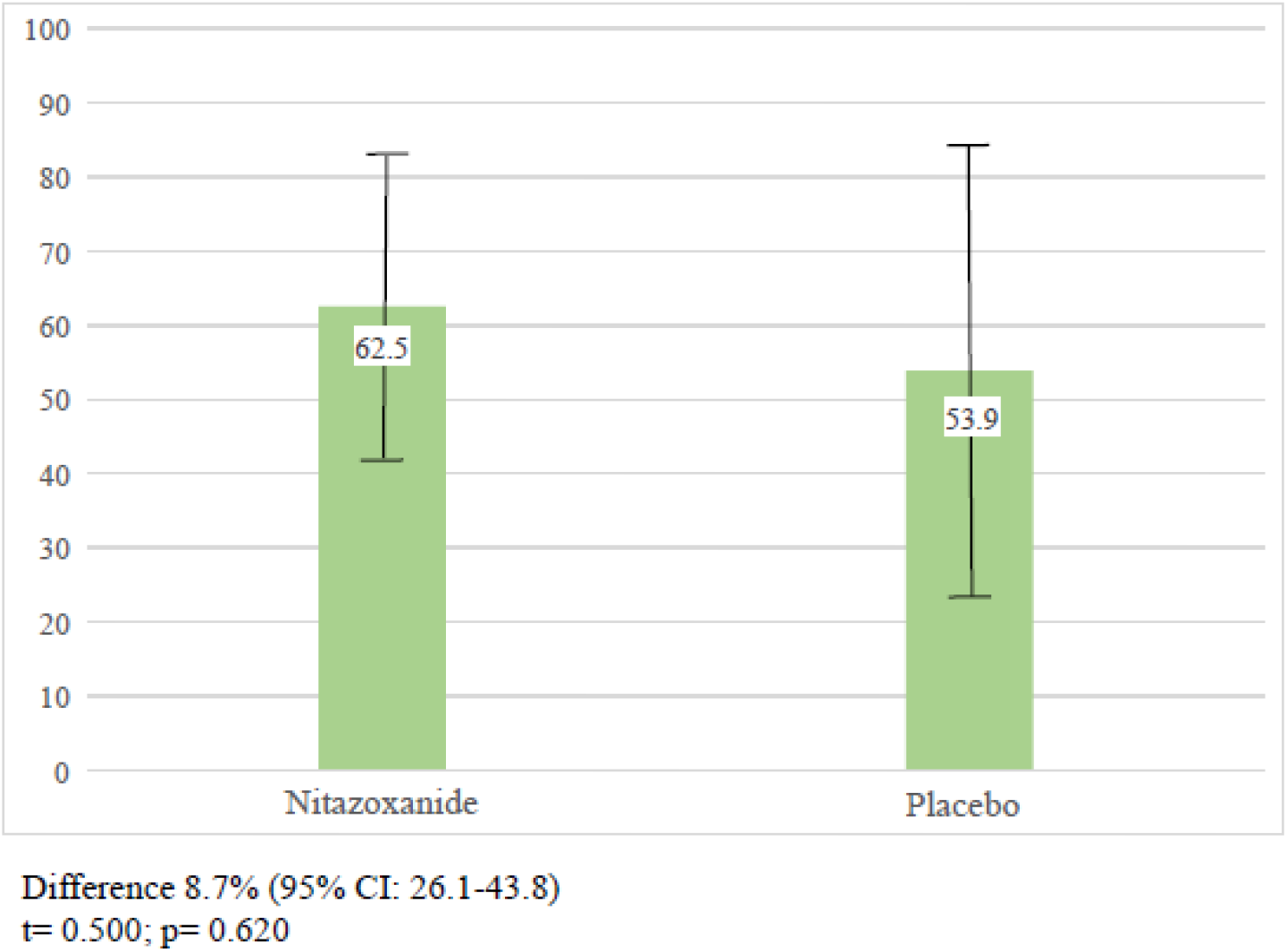
Primary efficacy results such as negativization of the viral load (N1 VL <100)Per treatment arm.Per-protocal analysis.

The primary efficacy analysis was also performed by intention-to-treat. In the case of patients with missing data, the Last Observation Carried Forward (LOCF) technique was used, which consists of assigning the last observation recorded.

A total of 40 patients were included in this analysis, 27 into the Nitazoxanide arm and 13 into the placebo arm. For all measurements, i.e., N1 Cq, absolute and relative variation in the viral load, N1 VL log, and absolute and relative variation in N1 VL log, a significant variation was observed between days 1 and 7, regardless of the treatment received, although the NTX group tends to show a greater variation.

The only discordant result in the intention-to-treat analysis compared to the per-protocol set is the N1 Cq increase ≥ 35%. Table 5 shows that 70.4% of the NTX patients achieved this result, compared to 53.8% of the patients who received placebo. This difference (16.6%; 95% CI: 16.5, 49.5) is not statistically significant (*t* = 1.014; *p* = 0.317), although the observed trend shows a higher proportion of patients with a variation ≥35% in the NTX arm.

**Table 5.** Secondary efficacy results (Patients with a variation ≥35% in N1 Cq) by treatment arm. Intention-to-treat analysis

| N1 Cq | Nitazoxanide<br>n=27 |  | Placebo<br>n=13 |  | Total<br>n=40 |  |
| --- | --- | --- | --- | --- | --- | --- |
|  | n | % | n | % | n | % |
| Day 7 | 19 | 70.4 | 7 | 53.8 | 26 | 65.0 |

Difference 16.6% (95% CI: -16.5, 49.5) t= 1.014; p= 0.317

For secondary efficacy results, including N1 Cq variation between Day 1 and Day 14, absolute and relative variation in the viral load, N1 VL log, and absolute and relative variation in N1 VL log, a significant variation was observed between days 1 and 14, regardless of the treatment received, although the NTX group tends to show a greater variation.

Regarding the safety of the treatment, GI adverse events were recorded in 26% of the patients treated with NTX compared to 15% with placebo, without significant differences between the two groups. Two deaths were recorded, one in each treatment arm. In both cases, the patients were older than 65 years and had other comorbidities.

## Discussion

We present here the interim analysis results of a randomized, placebo-controlled, single-blind, parallel group, pilot study to evaluate the efficacy and safety of NTX 500 mg QID for 14 days in patients with mild to moderate COVID-19 infection.

Viral load (VL) was select as the primary endpoint for being highly significant. In mild to moderate patients, clinical scales (such as the WHO COVID-19 ordinal scale) fail to display a great variation among patients; therefore, the use of such endpoints could impair the retrieval of meaningful differences. Furthermore, if we aim to prove the antiviral efficacy of a drug, it should be mandatory to use a virologic endpoint. As an example of the importance of using the VL as an endpoint, Chen et al. *(Chen et al., 2021)*, recently published a study where patients with higher viral load had a greater proportion of hospitalization rate than those with a better viral RNA clearance at day 7. These results were consistent with the finding that patients with delayed viral clearance had a more severe disease, suggesting that viral-load reducing agents might decrease rates of hospitalization.

A similar article *(Pujadas et al., 2020)* concluded that VL in COVID-19 correlate with disease phenotype, morbidity, and mortality, suggesting that it could also might modify isolation measures, since infectivity appears to be reduced in patients with low VL. Similarly, another article *(Riediker et al., 2020)* provides evidence of the association between viral load and the probability of infections, showing that higher viral loads correlate with greater probabilities of infections by the so-called superspreaders. Finally, another study by Zheng *(Zheng et al., 2020)* suggests that reducing viral loads by clinical means and strengthening the disease management during each stage of the severe disease should help prevent the spread of the virus.

In summary, early negativization or reduction of the viral load can potentially relieve the severity of this disease, resulting in a more controlled health system response avoiding collapse of the healthcare system.

Results from this small group of patients showed that NTX tended to reduce the viral load (VL) compared to placebo, and presented a favorable statistically significant difference in the number of patients with at least a 35% reduction in SARS-CoV-2 viral load at day 7 compared to placebo in patients with mild to moderate COVID-19.

We were not able to find differences among clinical endpoints because changes in the clinical parameters of mild to moderate patients are difficult to observe in small cohorts of patients. Based on the interim analysis of this pilot study, we were able to demonstrate NTX antiviral activity against COVID-19 in patients with mild to moderate infection. We believe this is a relevant finding, which proves previously reported NTX antiviral activity in pre-clinical grounds now in a clinical setting. Larger studies are needed to better understand the clinical impact of these findings. Moreover, final analysis of this and other currently ongoing studies are essential to provide complementary information about the most appropriate regimen in these patients.

Having low-cost, safe drug armamentarium, with proven anti-viral efficacy for the early treatment of COVID-19 infected patients is a goal that remains to be achieved. Studies like this, with well stablished virologic endpoints will help to define stronger strategies.

## Data Availability

All data referred to in the manuscript is available upon request

https://clinicaltrials.gov/ct2/show/NCT04463264?term=Nitazoxanide&cond=Covid19&cntry=AR&draw=2&rank=1

## Acknowledgements

The authors thank

Matias Tisi Baña ^1^ and Federico Piñero^1^. Protocol design and Natalia Alois ^1^. Virologic lab technician.

Pablo Liuboschitz and Eduardo Pirotzky Director and Medical Consultant respectively from Research & Development RA S.A. Clinical coordinating and monitoring of the study.

Ezequiel Klimovsky and Andrea A. Federico from K&H Latam Consulting SAS. Statistical analysis and collaboration in the writing of the manuscript.

---

**Annex 1.**

| N1 VL log | Nitazoxanide<br>n=23 |  | Placebo<br>n=13 |  | Total<br>n=36 |  |
| --- | --- | --- | --- | --- | --- | --- |
|  | Mean | SD | Mean | SD | Mean | SD |
| Day 1 | 4.2 | 1.6 | 3.6 | 1.7 | 4.0 | 1.7 |
| Day 7 | 1.9 | 1.1 | 2.0 | 1.3 | 1.9 | 1.2 |

| N1 VL log | Nitazoxanide<br>n=23 |  | Placebo<br>n=13 |  | Dif. | p | 95% CI of the difference |  |
| --- | --- | --- | --- | --- | --- | --- | --- | --- |
|  | Mean | SD | Mean | SD |  |  |  |  |
| Abs. Variation | -2.3 | 1.4 | -1.6 | 1.4 | 0.7 | 0.151 | -1.7 | 0.3 |
| Rel. variation | -53.8 | 27.1 | -43.0 | 28.5 | 10.8 | 0.269 | -8.7 | 30.2 |

Secondary efficacy results (Patients with a variation $\geq 35\%$ in N1 VL log) by treatment arm. Per-protocol analysis
| N1 VL log | Nitazoxanide<br>n=23 |  | Placebo<br>n=13 |  | Total<br>n=36 |  |
| --- | --- | --- | --- | --- | --- | --- |
|  | n | % | n | % | n | % |
| Day 7 | 18 | 78.3 | 7 | 53.8 | 25 | 69.4 |

Difference 24.5% (95% CI: 10.7 – 59.5)
t= 1.448; p= 0.162

